# Is the impact of social distancing on coronavirus growth rates effective across different settings? A non-parametric and local regression approach to test and compare the growth rate

**DOI:** 10.1101/2020.04.07.20049049

**Authors:** Neil Lancastle

## Abstract

Epidemiologists use mathematical models to predict epidemic trends, and these results are inherently uncertain when parameters are unknown or changing. In other contexts, such as climate, modellers use multi-model ensembles to inform their decision-making: when forecasts align, modellers can be more certain. This paper looks at a sub-set of alternative epidemiological models that focus on the growth rate, and it cautions against relying on the method proposed in (Pike & Saini, 2020): relying on the data for China to calculate future trajectories is likely to be subject to overfitting, a common problem in financial and economic modelling. This paper finds, surprisingly, that the data for China are double-exponential, not exponential; and that different countries are showing a range of different trajectories. The paper proposes using non-parametric and local regression methods to support epidemiologists and policymakers in assessing the relative effectiveness of social distancing policies. All works contained herein are provided free to use worldwide by the author under [CC BY 2.0](https://creativecommons.org/licenses/by/2.0/).

## Background

Mathematical models are routinely used in ‘prediction of epidemic trends’ (Pellis et al., 2020). The inherent uncertainty in these models, and in the early stages of an epidemic, is evident from the calls from epidemiologists at Oxford, UK, for ‘large scale serological surveys’ (Lourenço et al., 2020) to improve the accuracy of parameters such as susceptibility; and in the range of scenarios presented by epidemiologists at Imperial, UK (Ferguson et al., 2020). This difficult in choosing and parameterising the most effective prediction model is not unique to epidemiology. To resolve this, climate modellers use multi-model ensembles in their decision-making processes, acknowledging that ‘even if the perfect climate model did exist, any projection is always conditional on the scenario considered’ (Tebaldi & Knutti, 2007). Similarly, after the 2008 global financial crisis, over-reliance on equilibrium models by policymakers was recognised as both ‘ubiquitous’ (Bezemer, 2010) and ‘flawed’ (Bieta et al., 2012); leading to widespread calls for alternative approaches.

## Motivation

All models need to meet some basic criteria: they need to be theoretically grounded, and they need to be replicable across different settings, preferably with out-of-sample back-testing. This paper is motivated by a plethora of articles that focus on the ‘doubling rate’, which is the time taken for the total number of people infected to double. The ‘doubling rate’ is related to the growth rate (see **Table 2**) and successful reduction in the growth rate, by both individual and public health measures, is the goal of social distancing and other interventions. The aim is to flatten the curve and avoid a huge peak that would overwhelm health services (Wighton, 2020). The recent spate of articles includes contributions from non-epidemiologists and the media (Cuffe & Jeavans, 2020; Financial Times, 2020; Pike & Saini, 2020). The author is not arguing for alternative approaches to be given equal weight in the decision-making process, but to inform the debate by helping explain differences in outcomes between countries. In addition, this research is motivated by a desire to test the validity of assumptions made by Pike and Saini (2020), who used data from China to predict out of sample death rates in other countries after the implementation of social distancing.

## Ethics

There is a well-established principle in econometrics that using a limited dataset to forecast is generally frowned upon as overfitting (Bailey et al., 2016; Chalana, 2017). For that reason alone, the author would caution against relying too heavily on the data from China. Following a well-established precautionary principle that, in medical research, ‘one should take reasonable measures to avoid threats that are serious and plausible’ (Resnik, 2004), the author has refrained from using recent data on death statistics published by individual countries, and has instead focused on infection statistics (CSSE, 2020; UK-Government, 2020). Forecasting health care needs is known to be uncertain (Grenfell et al., 1994; Rein et al., 2011) but infection rates have the advantage that they are a leading indicator for pressure on intensive care beds. The author is required to get approval from his University ethics committee for research that ‘would induce psychological stress, anxiety or humiliation’ (DMU, 2020) and, given the author is not an epidemiologist, full approval and peer review were sought before pre-publication. Underestimates of intensive care beds could lead to a reduction in compliance with social distancing which, in turn, could lead increased harm to those most at risk of infection. Conversely, overestimates of intensive care beds could lead to unnecessary public health measures which, in turn, could lead to additional financial and economic harm.

## Research question and data

### Is the impact of social distancing on coronavirus growth rates comparable across different settings

Following the precautionary principle above, the null hypothesis proposed is that social distancing in different contexts is likely to lead to different outcomes. This is to avoid a Type 1 error – the rejection of a true null hypothesis – the equivalent of convicting an innocent person in a criminal trial. Historical data of cases in China are used. The start date for country comparisons is when cases reach fifty or more; information on relative lockdown dates is also shown in **Appendix 1**. The data were taken from the Humanitarian Data Exchange (Humanitarian Data Exchange, 2020) and, prior to 22^nd^ January 2020 for China, from China’s National Health Commission (National Health Commission, 2020). The R scripts used to run the tests are shown in **Appendix 2**. All works contained herein are provided free to use worldwide by the author under [CC BY 2.0](https://creativecommons.org/licenses/by/2.0/)

## Method

The first step in time series analysis is to collect and cleanse the data. This process requires transparency regarding how the data have been collected and handled, with any discrepancies resolved before analysis (DHSC, 2020). In finance and economics, the method most used is to winsorize to a stated range or percentage (Kroszner et al., 2007). Given the growth rate in infections for China fell below 0.005% after 45 days, data after this date are excluded. The author also excluded data from February 12th, when it was reported that infections in China jumped due to a change in the measurement method (Feuer, 2020) Model fitting using the full data is shown in **Appendix 4** as a robustness check.

An assumption that growth is exponential, in the early stages of an epidemic, is supported by the literature (Chowell et al., 2015; Nishiura et al., 2010), although the author is aware that other approximations exist, including polynomial. This paper does not attempt to review the theory behind this. For exponential-type growth, there are three choice of mathematical models: exponential-unrestrained; log-log (as per exponential, but with a longer period of decline); and logistic (approximately exponential at first, but with a reduced rate as an upper limit is approached). A logistic model fails the precautionary principle in that the carrying capacity, or c, can only be estimated ex-post. Hence, a logistic model is discarded.

The models being tested are therefore:

*Model 1 –ln(active cases) v time: y*= *ae*^*kt*^*or ln*(*y*)= *a*′+ *kt*

*Model 2 - ln(active cases) v ln(time):* ln(*y*)= *a* +*bln*(*t*)

*Where y = active* ^*1*^*cases; t = time; and a, a*′*and b are constants*

In financial time series analysis, for a stochastic (i.e.: random walk) process such as stock price movements, the rate of change in prices is generally used 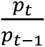: this technique is also applied here as 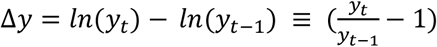, as a way to model when there are no new cases and the rate of change falls to zero 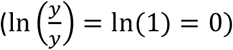. Hence, two further models are tested, namely:

*Model 3 – daily rate of change:* 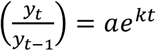 *or* Δ*y* = *a*′ + *kt*

*Model 4 - daily rate of change vs ln(time):* Δ*y*= *a*′+*bln*(*t*)

Next, *ln* (Δ*y*), a double-exponential model, is tested^2^. In this model, as the rate of change approaches zero (*y*_*t*_→*y*_*t*−1_)then (ln(Δ*y*) → − ∞).For example, when the daily change in new cases falls to 1%, ln(Δ*y*) is -4.6. Finally, the rate of change in the rate of change is included, or 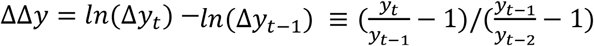. If there is an increase in the daily growth rate, this number is positive; if there is a decrease in the daily rate, this number is negative. It is not quite a stochastic process, because there should be a decrease in the daily growth rate over time, as the number of recovered cases increases:

*Model 5 – double-exponential: ln*(Δ*y*)= *a*′+ *kt*

*Model 6 – rate of rate of change:* Δ*yy*= *a*′+ *kt*

The next step in time series forecasting, after data cleansing, is to analyse the goodness-of-fit to alternative models. This is done graphically, by plotting time (x axis) against each model – the closer the result is to a straight line, the better the goodness-of-fit.

**Figure 1** plots the results for winsorized data from China. These confirm that Model 3 and Model 5 are an approximate linear fit, and Model 6 shows convergence towards zero. The author understands that log(time) relationships are found in nature (Dahly, 2017) as are hyper and double-exponential (Varfolomeyev & Gurevich, 2001). However, for the next part of the paper, the focus of the analyses is on relative growth rates, as shown in Model 5, and on falls/increases in the rate of change, as shown in Model 6.

**Figure 1:**
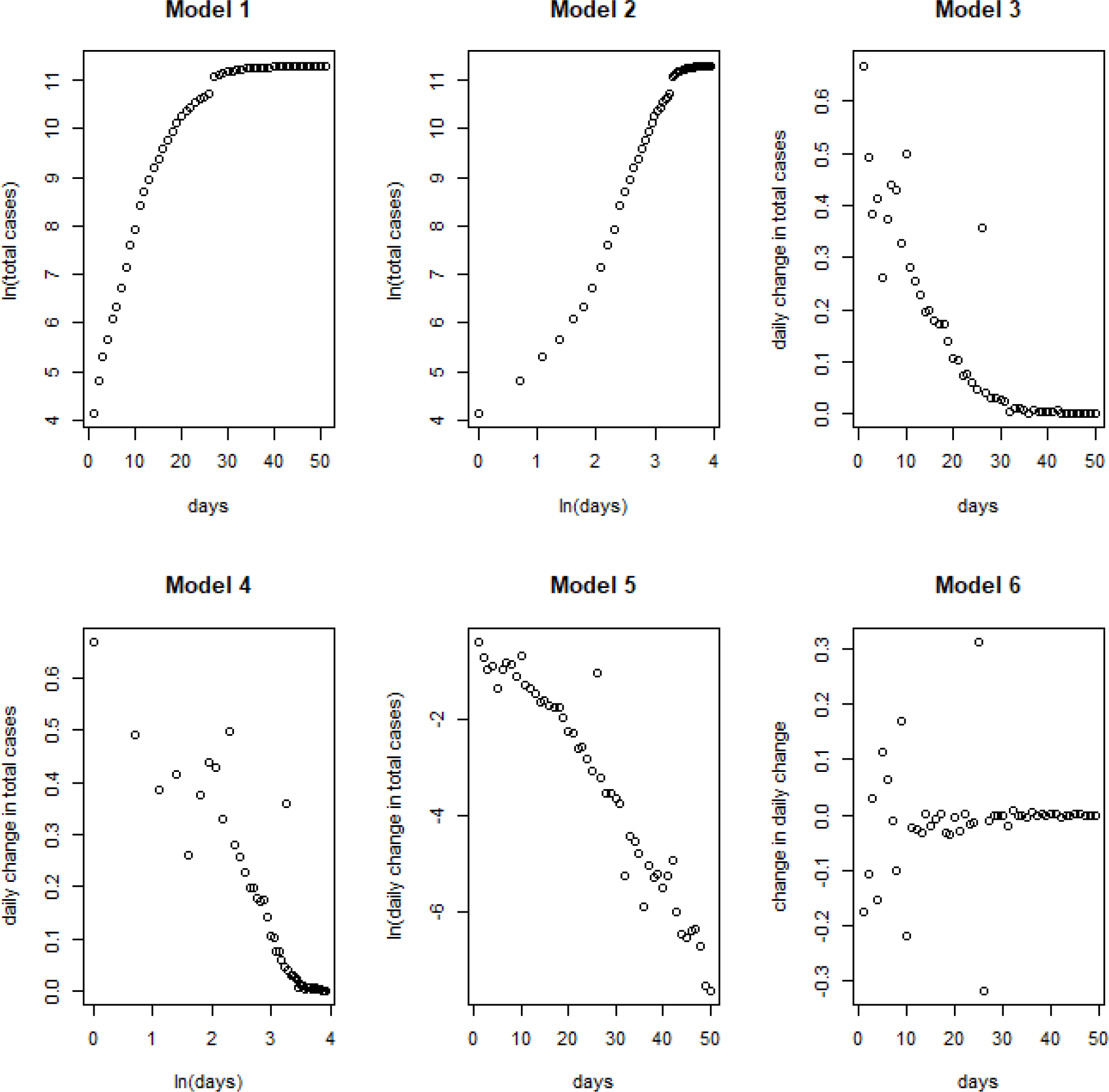
Model fitting.

We would not expect the data to be suitable for OLS regression, given there is convergence towards zero. This is confirmed by the descriptive statistics and Kolmogorov–Smirnov test in **Table 1**:

**Table 1:** Descriptive statistics.

| Time Series | Mean | Median | Skew | Kurtosis | KS Test |
| --- | --- | --- | --- | --- | --- |
| $\Delta y$ | 0.143 | 0.053 | 1.137 | 3.312 | Reject |
| $\ln(\Delta y)$ | -3.340 | -2.948 | -0.365 | 1.792 | Reject |
| $\Delta yy$ | -0.014 | -0.002 | -0.058 | 8.814 | Reject |

Therefore, not only should the reader be wary of overfitting using the China data to extrapolate; the reader should also be wary of overfitting via OLS (linear) regression.

The next part of this analysis applies a non-parametric method to determine how the growth rate varies vary over time. Data for China were split into four groups: 1-10 days; 11-20 days; 21-30 days, and 31-40 days. From a Kruskal-Wallis test, followed by pairwise comparisons using Wilcoxon rank sum test (results not shown) the growth rates in each period are significantly different. To illustrate this, **Appendix 6** shows locally estimated scatterplot smoothing (LOESS) for the first three groups - 1-10 days; 11-20 days; and 21- 30 days – using Model 5. For comparison, the author understands that the paper by (Pike & Saini, 2020) uses Model 3.

**Appendix 6** show a marked drop in the growth rate over 30 days, with most of the drop occurring in the second period – 11-20 days. These results provide researchers with some visible patterns that might, or might not, emerge from the data for other countries as social distancing continues. Hence, the last part of this analysis uses Model 5 to compare all countries. From **Table 2**, when the number of new cases approaches zero, the model approaches −∞; alternatively, when the number of new cases falls to 1%, the model is approximately -4.6, which is a doubling rate of every 70 days or so. Hence, the lower the y-axis the better.

**Table 2:** Interpreting Figures 2-5.

| Total Cases | % daily increase | Y-axis | Growth rate |
| --- | --- | --- | --- |
| 100 | | $\ln^3$ | |
| 200 | 100% | -0.37 | Doubling every day |
| 400 | 100% | -0.37 |  |
| 600 | 50% | -0.90 | Doubling in less than two days |
| 800 | 33% | -1.25 | Doubling less than three days |
| 1000 | 25% | -1.50 | Doubling in less than four days |
| 1010 | 1% | -4.61 | Doubling in about 70 days |
| 1011 | 0.1% | -6.92 | Doubling in more than 500 days |

The following figures are organized as follows. **Figure 2** show growth rates for the fifty countries with the highest total cases (in descending order). The x-axis starts from the day that cases reach fifty in that country; China is the only country to have dipped below a 1% daily growth rate; although Austria, South Korea and Norway have had a similar trajectory so far.

**Figure 2.**
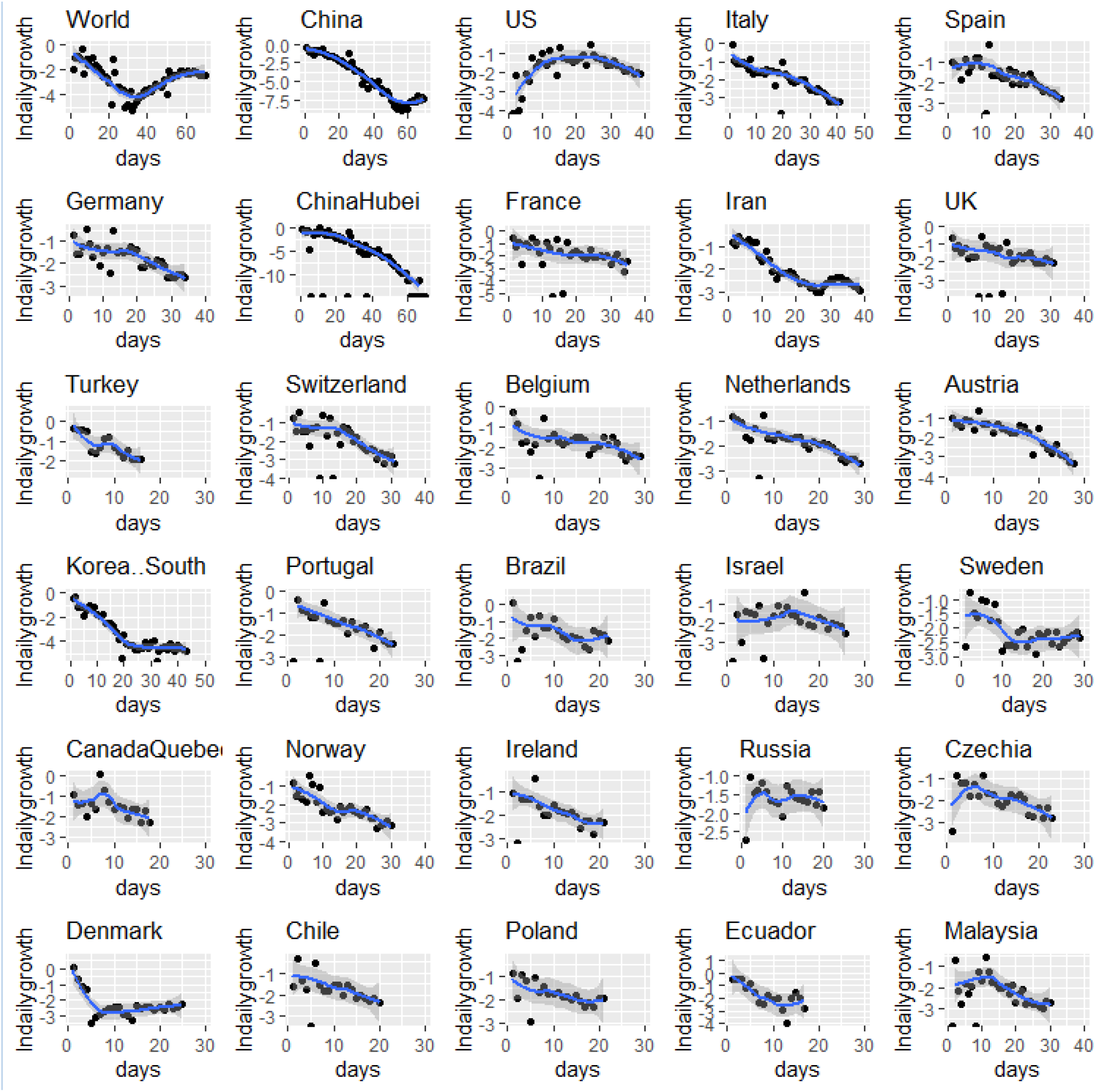
Growth rates for highest total cases, as at 7^th^ April 2020.

**Figure 3** focusses on the highest median daily growth rates in the first period - 1-10 days - after a country or region has reached fifty cases. The countries with the highest initial growth rates are Iran, Turkey, Italy, South Korea and Spain; Cameroon is also included in this group because it has the highest median growth rate in the second period - 11-20 days.

**Figure 3.**
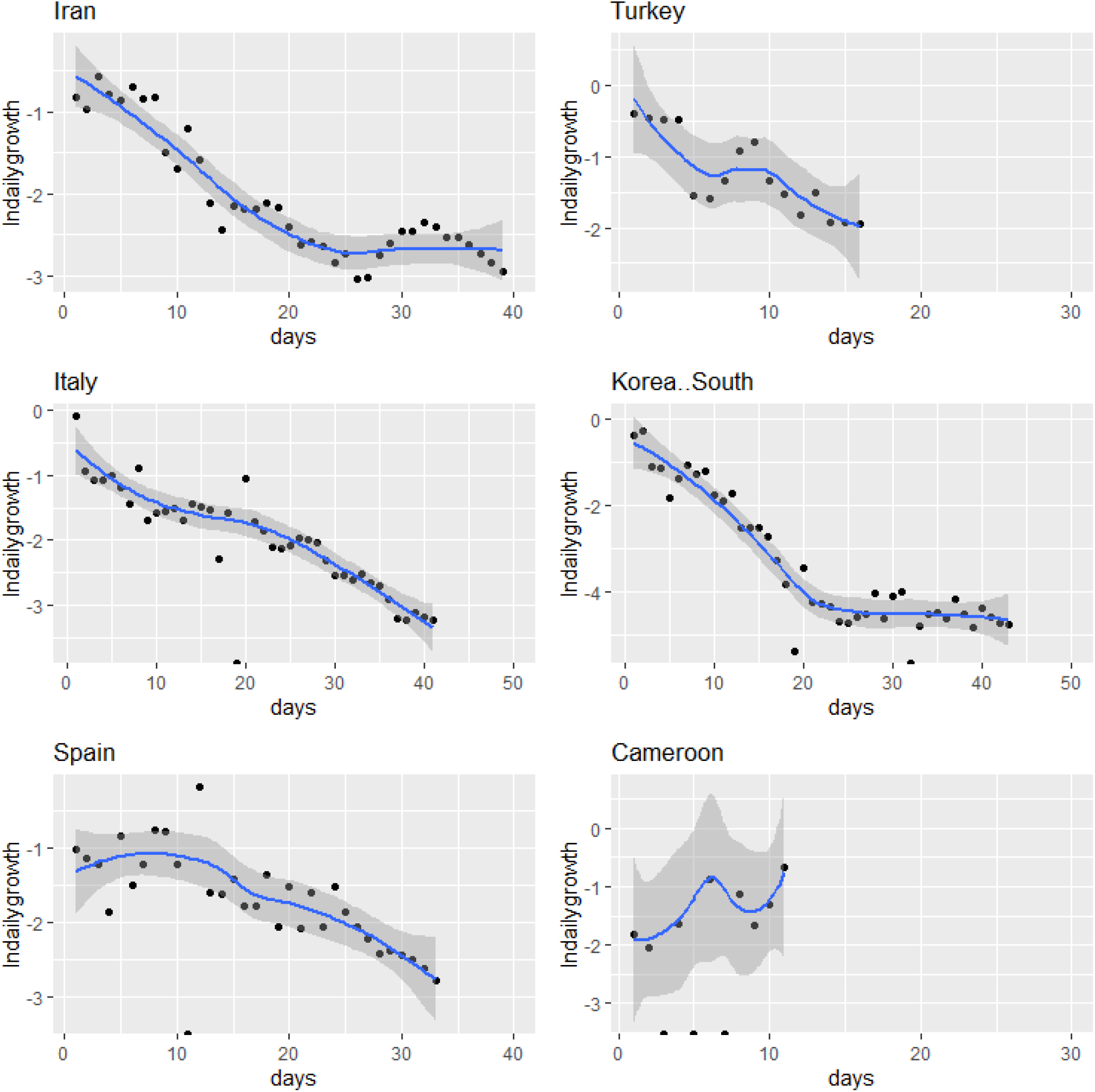
Highest median growth rates in first 1-10 days (Cameroon on day 11)

**Figure 4** focusses on those countries and regions where growth rates are lowest in the third period – 21-30 days. With the caveats, above, about data and methodological issues, these are countries that have managed to bring the pandemic under control within 30 days. Eleven regions of China, that would dominate this table, have been excluded so that other low growth rate countries are revealed: South Korea, Lebanon, Australia NSW, Bahrain, and British Columbia. Lastly, **Figure 5** shows those countries where growth rates have fallen most compared to earlier period; this includes countries that had high initial growth rates but are succeeding in bringing down the growth rate quickly – the mean rate of rate of change is most negative in these countries - France, Spain, Switzerland, Italy, UK and Norway.

**Figure 4.**
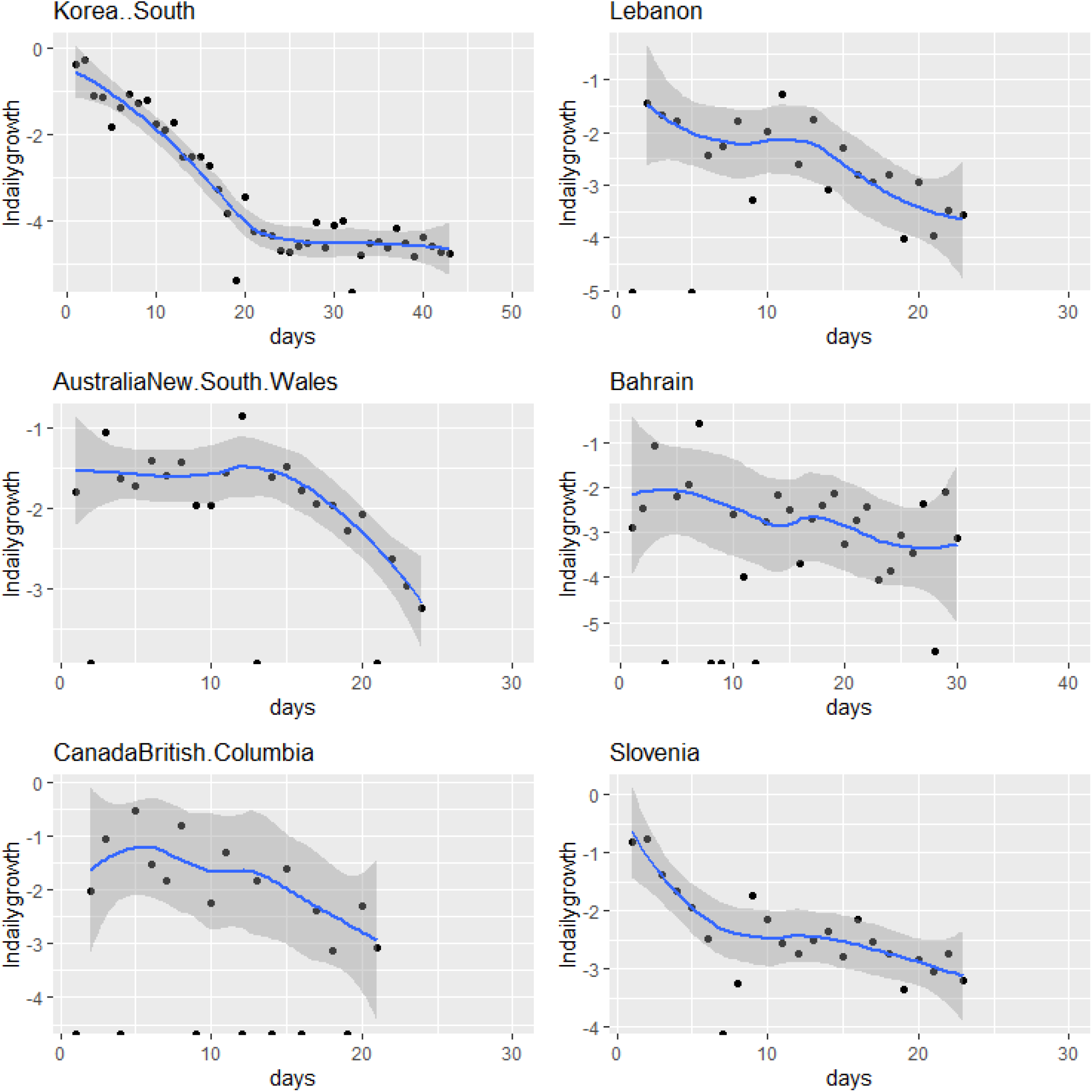
Lowest median growth rates in period 21-30 days (excluding China)

**Figure 5.**
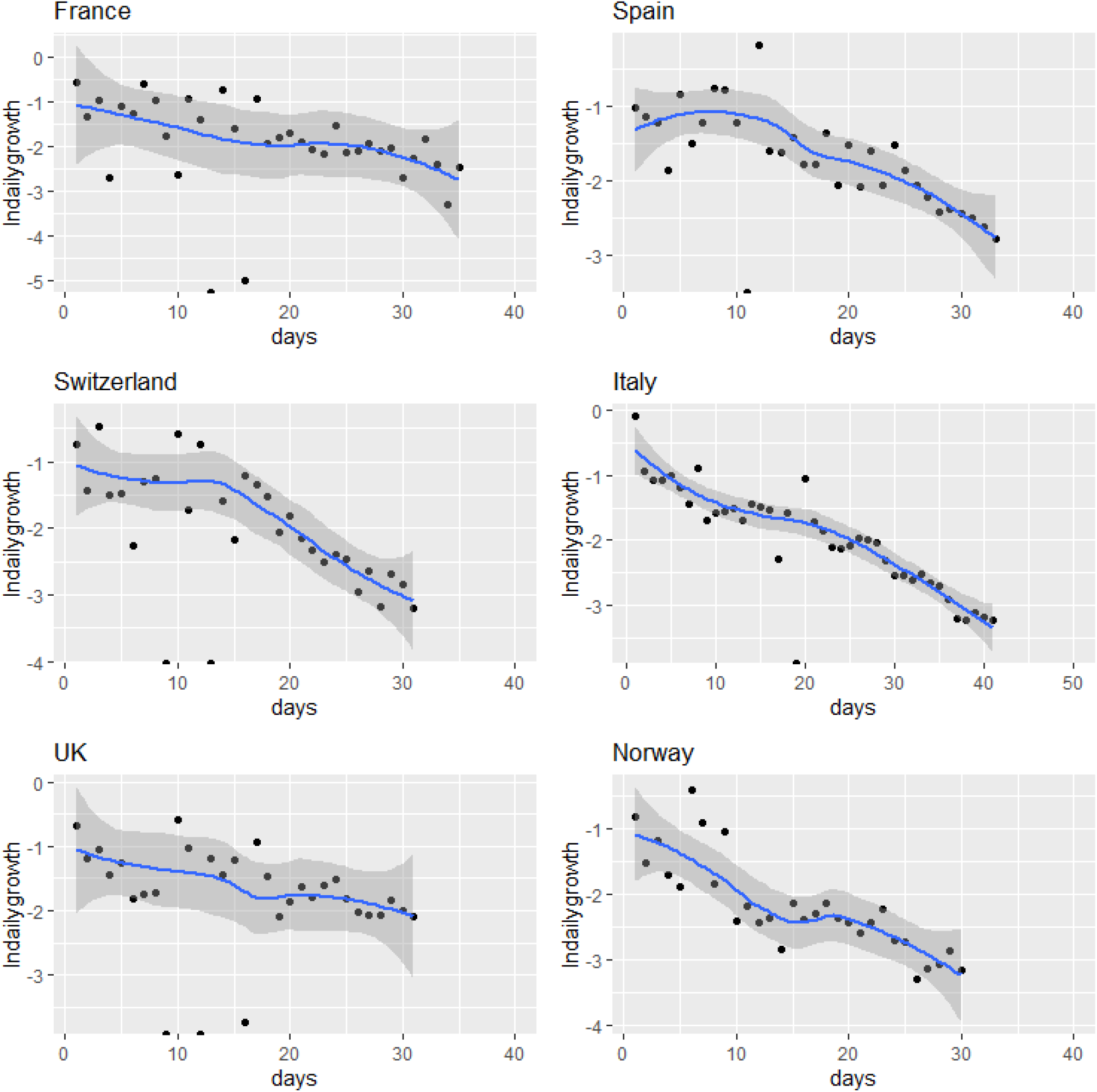
Lowest mean fall in growth rates from 1-30 days.

## Conclusion

The methodology presented here is a non-parametric and graphical approach, using freely available software, to test whether different phases of social distancing are comparable across different settings. If the null hypothesis, that social distancing is not comparable, can be rejected, then policymakers can be reassured that their social distancing measures are being successful. All works contained herein are provided free to use worldwide by the author under [CC BY 2.0](https://creativecommons.org/licenses/by/2.0/)

With the above caveats above data quality, transparency, and consistency, the data for China show that strong social distancing led initially to a high reduction in the growth rate; then a rapid decline towards a daily growth rate of <1% by 30 days; followed by a stable period of low transmission. There is some evidence, from the data shown in **Figures 2-5**, that European countries in the panel are showing similar trajectories since social distancing. However, China dominates the results for **Figure 4** – lowest median growth rates after 21-30 days – and only a handful of countries are showing similar reductions.

Whilst there are lower initial growth rates in several countries, such as Bahrain, Iran, Kuwait, Qatar and Singapore – the evidence presented thus far supports the null hypothesis that social distancing in different contexts is likely to lead to different outcomes. To the author’s knowledge, there is no theoretical basis for assuming that other countries will follow China’s trajectory. Rather, the evidence, from the results presented, is that a range of different outcomes are likely. Policymakers will need to take account of differences in social, cultural, political, and economic factors; to take account of different data collection, quality, and consistency issues across different settings; and should take account of the lag, of around 10 days, between the start of social distancing and any significant impact on the number of cases. In other words, policy needs to be based on what is happening in hospitals and communities.

These are tentative conclusions, and the results require replication and confirmation – this is a pre-print of the paper. With these caveats; this approach could inform the policy debate around the effectiveness of different approaches of social distancing. There are three parts to the methodology: i) determining if data from different countries shows a similar exponential distribution using Models 5 and Model 6 ii) using graphical and LOESS approaches to determine whether the data from different contexts are following the same trajectories in equivalent periods 1-10, 11-20 and 21-30 days, and iii) using non-parametric approaches to see if growth rates are significantly lower from one period to the next.

In terms of prediction, the author supports those epidemiologists who are calling for better data (Lourenço et al., 2020) through testing, and who are presenting a range of scenarios (Ferguson et al., 2020); until such time that a consensus emerges from a multi-modelling perspective. In particular, the author cautions against giving much weight to models that extrapolate from growth rates, or that assume other countries will necessarily follow the same path as China, in order to avoid a type 1 error and rejection of a true null hypothesis. To paraphrase the infinite monkey theorem, that is not to say that, with enough predictive models, at least one of them will fit the available data *by chance*. Following the precautionary principle, the author recommends against policy recommendations that are not supported by clinical observations in the field and/or by a convergence of predictions from different epidemiological prediction models, such as the SIR models at Oxford, UK and Imperial, UK.

Lastly, as well as using this method to compare different phases of social distancing across countries, the methodology might also be useful for regional analyses; and for sociologists, economists and political scientists to determine the social, economic, and political factors that underpin successful social distancing.

## Data Availability

The data were taken from the Humanitarian Data Exchange (Humanitarian Data Exchange, 2020) and, prior to 22nd January 2020 for China, from China’s National Health Commission (National Health Commission, 2020). The R scripts used to run the tests are shown in Appendix 2. All works contained herein are provided free to use worldwide by the author under CC BY 2.0

https://data.humdata.org/dataset/novel-coronavirus-2019-ncov-cases

## Acknowledgments

The author would like to thank Frank Kwabi, Dave Walsh, Tom Allen, Jonathan Rose, Linda Hickson, Olivia Lancastle, Fred Mear, Mark Kirstein, Stuart MacDonald and Louis Ellam for their useful comments.

## Appendix 1

~~~
# first day of lockdowns – day 0
# China 23rd January (24th January - most Wuhan provinces) - 830 cases
# UK Saturday 21st March (announced 5pm on Friday 20th but pubs still open) - 5018 cases
# Spain Sunday 15th March (announced on Saturday evening) - 7375 cases
# Italy Sunday 8th March (announced early in morning) - 7375 cases
# France Tuesday 17th March (announced on Monday to start next day) - 7730 cases
# US Tuesday 16th March (announced on Monday to start next day) - 4596 cases
~~~

## Appendix 2 R Scripts

~~~
######################################################################
# set up some useful packages
install.packages(“ moments”)
library(moments)
install.packages(“ ggplot2”)
library(“ ggplot2”)
install.packages(“ gridExtra”)
library(“ gridExtra”)
install.packages(“ ggfortify”)
library(ggfortify)
#######################################################################
# Read in the data from a csv file
alldata <- read.table(“ c:/data/allHUMdata.csv”, header = TRUE, sep = “,”)
attach(alldata)
x <- colnames(alldata)
# get the data for China
totalcases <- alldata[,3]
# Omit less than 50 cases, then truncate to 71 days
totalcases[totalcases < 50] <- NA
totalcases = na.omit(unclass(totalcases)) totalcases <- totalcases[1:71]
totalcases <- ts(totalcases)
# work out the difference, for Models 3 and 5
model3China = log(totalcases) - log(lag(totalcases,-1))
########################################################################
# FIGURE 1 Model fitting - ALL DATA UP TO 50 DAYS
par(mfrow=c(2,3))
totalcases = unclass(totalcases)
lntotalcases = log(totalcases)
plot(c(1:71), lntotalcases,
  xlab = “ days”, ylab=“ ln(total cases)”,
  main = “ Model 1”)
plot(log(c(1:71)), lntotalcases,
  xlab = “ ln(days)”, ylab=“ ln(total cases)”,
  main = “ Model 2”)
plot(c(1:70), model3China,
  xlab = “ days”, ylab=“ daily change in total cases”,
  main = “ Model 3”)
plot(log(c(1:70)), model3China,
  xlab = “ ln(days)”, ylab=“ daily change in total cases”,
  main = “ Model 4”)
plot(c(1:70), log(model3China),
  xlab = “ days”, ylab=“ ln(daily change in total cases)”,
  main = “ Model 5”)
model6China = model3China - lag(model3China,-1)
plot(c(1:69), model6China,
  xlab = “ days”, ylab=“ change in daily change”,
  main = “ Model 6”)
###############################################################################
# FIGURE 1 Model fitting - excluding February 12th
# get the winzorised data for China
totalcases <- alldata[,4]
# Omit less than 50 cases, then truncate to 51 days
totalcases[totalcases < 50] <- NA
totalcases = na.omit(unclass(totalcases))
totalcases <- totalcases[1:51]
totalcases <- ts(totalcases)
# work out the difference, for Model 3
model3China = log(totalcases) - log(lag(totalcases,-1))
par(mfrow=c(2,3))
totalcases = unclass(totalcases)
lntotalcases = log(totalcases)
plot(c(1:51), lntotalcases,
  xlab = “ days”, ylab=“ ln(total cases)”,
  main = “ Model 1”)
plot(log(c(1:51)), lntotalcases,
  xlab = “ ln(days)”, ylab=“ ln(total cases)”,
  main = “ Model 2”)
plot(c(1:50), model3China,
  xlab = “ days”, ylab=“ daily change in total cases”,
  main = “ Model 3”)
plot(log(c(1:50)), model3China,
  xlab = “ ln(days)”, ylab=“ daily change in total cases”,
  main = “ Model 4”)
plot(c(1:50), log(model3China),
  xlab = “ days”, ylab=“ ln(daily change in total cases)”,
  main = “ Model 5”)
model6China = model3China - lag(model3China,-1)
  plot(c(1:49), model6China,
  xlab = “ days”, ylab=“ change in daily change”,
main = “ Model 6”)
########################################################################
# Table 1 Descriptive statistics using winzorised data
x <- rnorm(1000)
# Daily change
summary(model3China)
skewness(model3China)
kurtosis(model3China)
# Do x and y come from the same distribution?
ks.test(model3China, x)
# ln(daily change)
summary(log(model3China))
skewness(log(model3China))
kurtosis(log(model3China))
# Do x and y come from the same distribution?
ks.test(log(model3China), x)
# rate of rate of change
summary(model6China)
skewness(model6China)
kurtosis(model6China)
# Do x and y come from the same distribution?
ks.test(model6China, x)
##########################################################################
# compare groups
alldata <- read.table(“ c:/data/chinakruskal.csv”, header = TRUE, sep = “,”)
attach(alldata)
kruskal.test(data ∼ group, data = alldata)
pairwise.wilcox.test(data, group,
      p.adjust.method = “ BH”)
############################################################################
# Plot LOESS for the first three groups, Model 5
# get the winzorised data for China again, just in case
# Read in the data from a csv file
alldata <- read.table(“ c:/data/allHUMdata.csv”, header = TRUE, sep = “,”)
attach(alldata)
x <- colnames(alldata)
# get the winzorised data for China
totalcases <- alldata[,4]
# Omit less than 50 cases, then truncate to 51 days
totalcases[totalcases < 50] <- NA
totalcases = na.omit(unclass(totalcases))
totalcases <- totalcases[1:51]
totalcases <- ts(totalcases)
# work out the difference, for Model 3
model3China = log(totalcases) - log(lag(totalcases,-1))
# Split the data into three groups
model3 = log(totalcases) - log(lag(totalcases,-1))
# get the first 30 days only
model3trunc <- model3[1:30]
# get the log, for Model 5
model5trunc = log(model3trunc)
#split into three ten-day periods (where available)
model5split <- split(model5trunc, ceiling(seq_along(model5trunc)/10))
# days 1-10
# create a data frame, for the LOESS plots
days <- c(1:10)
Y <- as.numeric(unlist(model5split[1]))
df <- data.frame(lndailychange = Y, days)
# then plot the results
p1 <- ggplot(df, aes(days, lndailychange)) + geom_point()+ ggtitle(“ 1-10 days”)+ geom_smooth(method
= “ loess”, level = 0.99)
# days 11-20
# create a data frame, for the LOESS plots
days <- c(11:20)
Y <- as.numeric(unlist(model5split[2]))
df <- data.frame(lndailychange = Y, days)
# then plot the results
p2 <- ggplot(df, aes(days, lndailychange)) + geom_point()+ ggtitle(“ 11-20 days”)+
geom_smooth(method = “ loess”, level = 0.99)
# days 21-30
# create a data frame, for the LOESS
plots days <- c(21:30)
Y <- as.numeric(unlist(model5split[3]))
df <- data.frame(lndailychange = Y, days)
# then plot the results
p3 <- ggplot(df, aes(days, lndailychange)) + geom_point()+ ggtitle(“ 21-30 days”)+
geom_smooth(method = “ loess”, level = 0.99)# + ylim(0,0.4)
grid.arrange(p1, p2, p3, ncol=3, top = “ Model 5 - China”)
#############################################################################
#################################################################
####### FULL DATA
# Read in csv file
alldata <- read.table(“ c:/data/allHUMdata.csv”, header = TRUE, sep = “,”)
attach(alldata)
x <- colnames(alldata)
# create an array for the results
column.names <- x
row.names <- c(“ country”, “ total cases”, “ max daily cases”, “ 1-10 days median daily growth (ln)”, “ 11-20
days median daily growth (ln)”, “ 21-30 days median daily growth (ln)”, “ 30 day median daily growth”,
“ 30 day mean daily fall in growth rate”)
results <- array(, c(103,8), dimnames=list(column.names, row.names))
n=102
# first series n=26 and loop (i in 2:n)
# second series n=51 and loop (i in 27:n)
# then get summary statistics for all of the series
# be sure to add all of the plots to p
p <- list()
# Loop over all columns, omit total <50
for(i in 2:n) {
   temp <- alldata[, i] # Get data for ith column and save in a new temporary object temp
      totalcases <- ts(temp)
   # first, total cases
   # start omit < 50
   totalcases[totalcases < 50] <- NA
   totalcases = na.omit(unclass(totalcases))
# end omit < 50
   totalcases <- ts(totalcases)
   results[i,1] = max(totalcases)
   # then, max daily cases
   dailycases = totalcases - lag(totalcases,-1)
   results[i,2] = max(dailycases)
   # Model 3 - median daily growth - ln
model3 = log(totalcases) - log(lag(totalcases,-1))
# get the first 30 days only
   model3trunc <- model3[1:30]
   # split into three ten-day periods (where available)
   model3split <- split(model3trunc, ceiling(seq_along(model3trunc)/10))
   # days 1-10
   res1 <- lapply(model3split[1],median,na.rm=TRUE)
   res1 <- as.numeric(unlist(res1))
   results[i,3] <- res1
   # days 11-20
   res2 <- lapply(model3split[2],median,na.rm=TRUE)
   res2 <- as.numeric(unlist(res2))
   results[i,4] <- res2
   # days 21-30
   res3 <- lapply(model3split[3],median,na.rm=TRUE)
   res3 <- as.numeric(unlist(res3))
   results[i,5] <- res3
###############################################
   # Model 3 regression
   #mod <- lm(log(totalcases) ∼ log(lag(totalcases,-1)))
   # TEST
   #x = c(1,2,3)
   #y = c(2,4,6)
   #mod <- lm(x ∼ y)
   #coeff <- coefficients(mod)
   #paste0(“ y = “, round(coeff[2],1), “ *x “, round(coeff[1],1))
   #mod <- lm(lag(totalcases,-1) ∼ totalcases)
   #coeff <- coefficients(mod)
   ##paste0(“ y = “, round(coeff[2],1), “ *x “, round(coeff[1],1))
   #results[i,6] <- paste0(“ y = “, round(coeff[2],6), “ *x “, round(coeff[1],6))
################################################
   # Model 5 - change in daily growth
   # force into a time series
   model5ts <- ts(log(model3), start(1), frequency=1)
   # plots as in pre-print
   #ts <- plot.ts(model5ts, xlab = “ Time”, main = column.names[i], ylab = “ change in daily growth”,
ylim=c(-6,0))
   # create a data frame, for the LOESS plots
   if (length(model5ts) < 30) {
      days <- c(1:30)
      Y <- model5ts[1:30]
      Y <- as.matrix(Y)
   } else if (length(model5ts) < 40) {
      days <- c(1:40)
      Y <- model5ts[1:40]
      Y <- as.matrix(Y)
   } else if (length(model5ts) < 50) {
         days <- c(1:50)
      Y <- model5ts[1:50]
      Y <- as.matrix(Y)
   } else if (length(model5ts) < 60) {
      days <- c(1:60)
      Y <- model5ts[1:60]
      Y <- as.matrix(Y)
   } else {
      days <- c(1:70)
      Y <- model5ts[1:70]
      Y <- as.matrix(Y)
   }
   df <- data.frame(lndailygrowth = Y, days)
   # then plot the results
   p1 <- ggplot(df, aes(days, lndailygrowth)) + geom_point()+ ggtitle(column.names[i])+
geom_smooth(method  = “ loess”, level = 0.99)
   p[[i]] <- p1
   # 30 day median
   model3trunc <- model3[1:30]
   res1 <- as.numeric(unlist(model3trunc))
   res1 <- sapply(res1,median,na.rm=TRUE)
   results[i,6] <- median(res1)
   # 30 day median drop in daily rate
   model3trunc <- model3[1:30]
   model3trunc <- ts(model3trunc, start(1), frequency=1)
   model3trunclag <- lag(model3trunc,-1)
   model3trunclag <- ts(model3trunclag, start(1), frequency=1)
   model3ts <- model3trunc -lag(model3trunclag,-1)
   res1 <- as.numeric(unlist(model3ts))
   results[i,7] <- median(res1)
   # 30 day median drop in daily rate
   model3trunc <- model3[1:30]
   model3trunc <- ts(model3trunc, start(1), frequency=1)
   model3trunclag <- lag(model3trunc,-1)
   model3trunclag <- ts(model3trunclag, start(1), frequency=1)
   model3ts <- model3trunc -lag(model3trunclag,-1)
   res1 <- as.numeric(unlist(model3ts))
   results[i,8] <- mean(res1)
###############################################
   # Model 5 regression
   # need to get rid of -Inf
   #mod <- lm(model5ts ∼ lag(model5ts,-1))
   #results[i,7] <- mod$coefficients[1]
################################################
   #install.packages(“ forecast”)
   #library(forecast)
   #auto.arima(model5ts, trace=TRUE)
   #model5 = model5ts - lag(model5ts,-1)
   #arimastats <- stats::arima(model5ts, order=c(1,0,0), method=“ ML”)
   #next20 <- predict(arimastats, 20)
   #results[i,6] <- log(median(next20$pred))
}
# show results
# Figure 2 - top 50
grobs = list(p[[2]], p[[3]],p[[5]],p[[6]],p[[7]],p[[8]],p[[9]],p[[10]],p[[11]],p[[12]],p[[13]],p[[14]],
p[[15]],p[[16]],p[[17]],p[[18]],p[[19]],p[[20]],p[[21]],p[[22]],p[[23]],p[[24]],p[[25]],p[[26]],p[[27]],p[[28]],
p[[29]],p[[30]],p[[31]],p[[32]])
do.call(grid.arrange,grobs)
# Figure 3 - highest median in days 1-10
grobs = list(p[[11]], p[[13]],p[[6]],p[[18]],p[[7]],p[[91]])
do.call(grid.arrange,grobs)
# Figure 4 - lowest median in days 1-10
grobs = list(p[[82]],p[[76]],p[[102]],p[[61]],p[[64]],p[[75]])
do.call(grid.arrange,grobs)
# Figure 5 - lowest median in days 21-30
grobs = list(p[[18]],p[[92]],p[[40]],p[[82]],p[[59]],p[[72]])
do.call(grid.arrange,grobs)
# Figure 5 - lowest mean fall in growth rates
grobs = list(p[[10]],p[[7]],p[[14]],p[[6]],p[[12]],p[[24]])
do.call(grid.arrange,grobs)
write.table(results, file=“ c:/data/allresults.csv”, append = FALSE, sep = “,”, dec = “ .”,
row.names = TRUE, col.names = TRUE)
#####################################################################################
###
~~~

## Appendix 3

**Figure 5:**
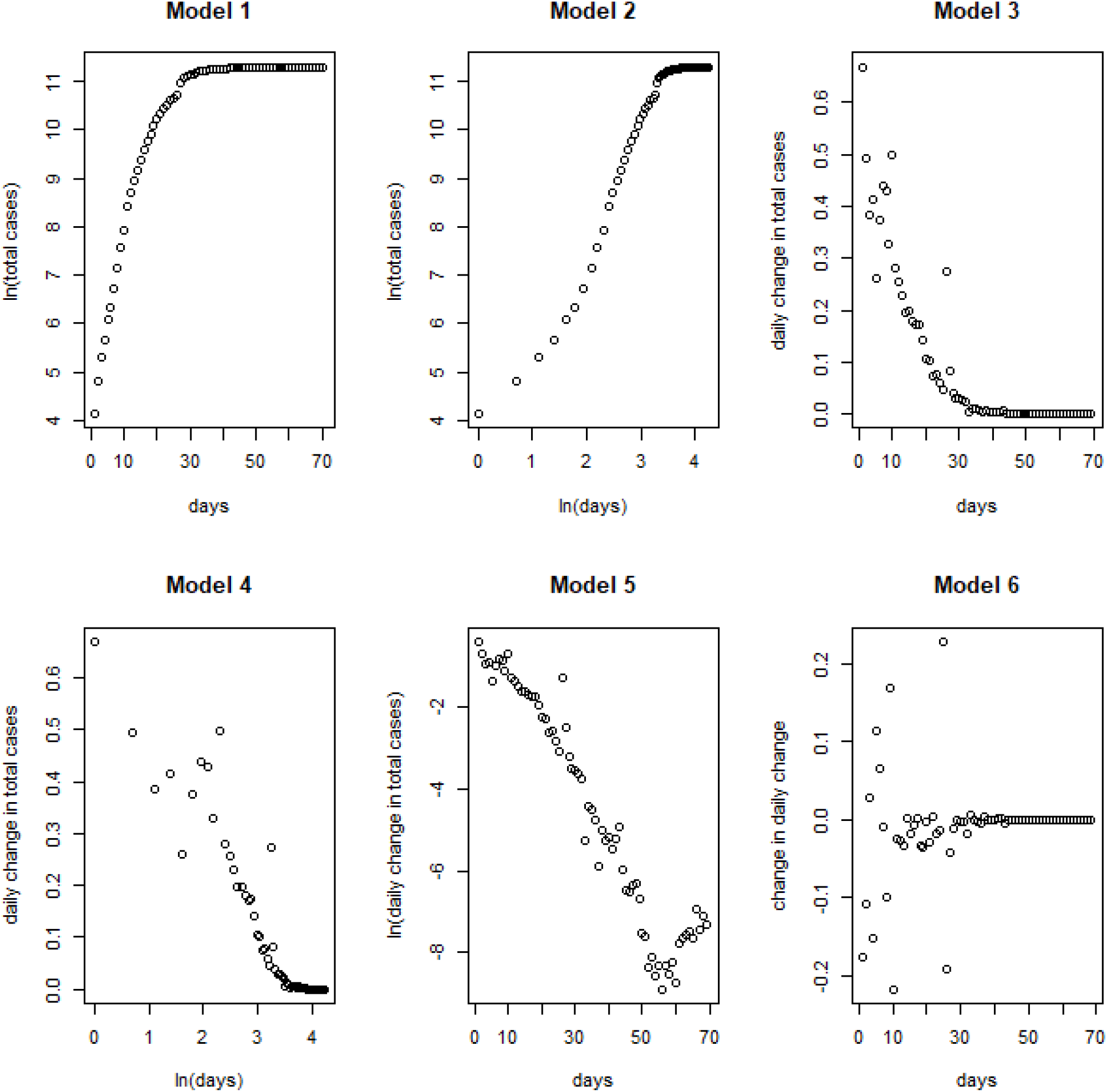
Model fitting – full data.

## Appendix 4 LOESS for China periods 1-10, 11-20 and 21-30 days

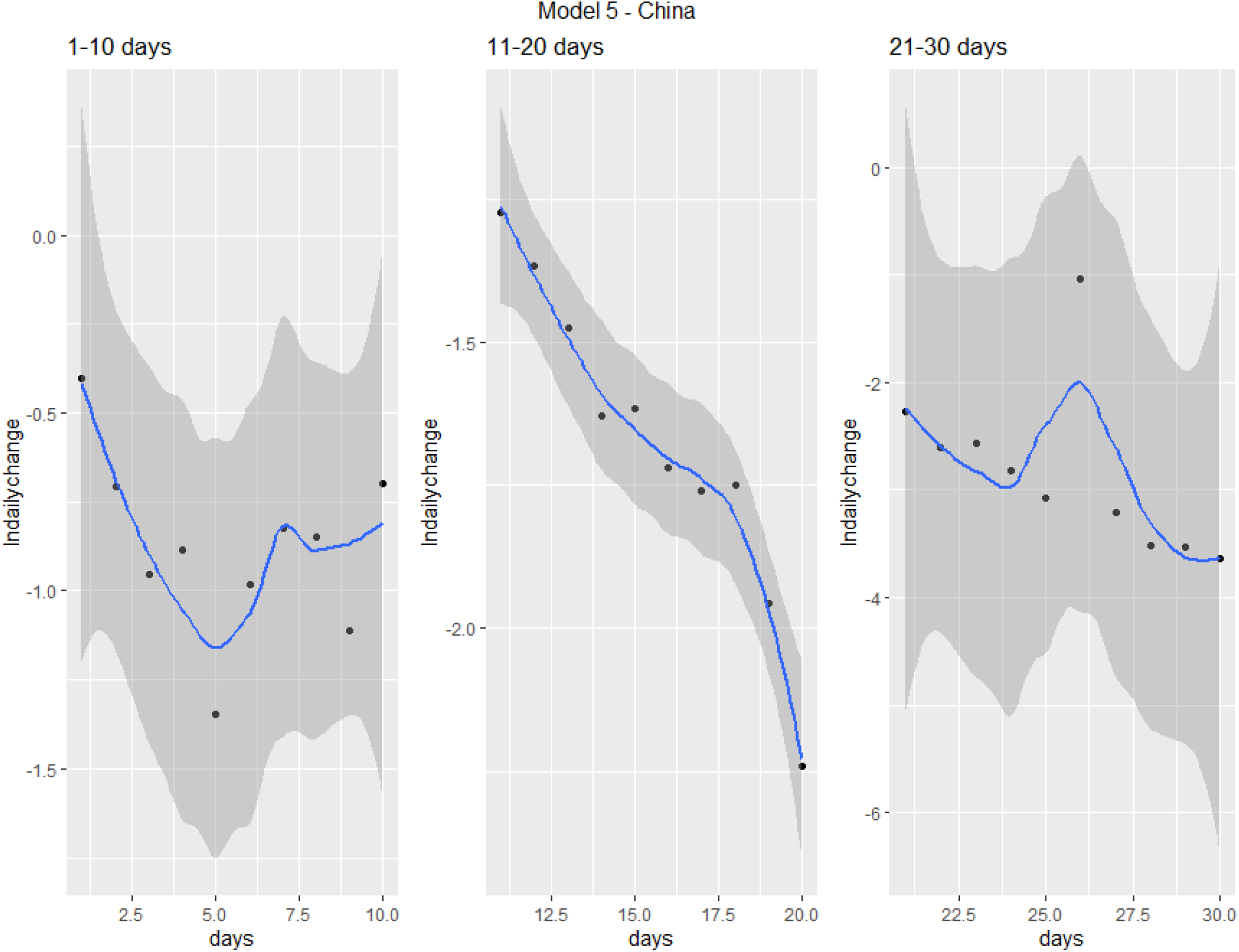

## Footnotes

1 For this paper, good data for recovered cases are not available. The author considered using new cases in the last 10 days as a proxy for active cases, but the analysis would be incomplete. Instead, total cases have been used. This is likely to bias results after 30 days or so, when recovered cases are likely to outweigh active cases.

2 The author understands this to be a Laplace distribution.

3 log in R

## Notes

### Competing Interest Statement

The authors have declared no competing interest.

### Funding Statement

No external funding

